# Multi-omic associations of epigenetic age acceleration are heterogeneously shaped by genetic and environmental influences

**DOI:** 10.1101/2024.06.28.24309638

**Authors:** Gabin Drouard, Sannimari Suhonen, Aino Heikkinen, Zhiyang Wang, Jaakko Kaprio, Miina Ollikainen

## Abstract

Connections between the multi-ome and epigenetic age acceleration (EAA), and especially whether these are influenced by genetic or environmental factors, remain underexplored. We therefore quantified associations between the multi-ome comprising four layers – the proteome, metabolome, external exposome, and lifestyle – with six different EAA estimates. Two twin cohorts were used in a discovery-replication scheme, comprising respectively young (N=642; mean age= 22.3) and older (N=354; mean age=62.3) twins. Within-pair twin designs were used to assess genetic and environmental effects on associations. We identified 40 multi-omic factors, of which 28 were proteins, associated with EAA in the young twins while adjusting for sex, smoking, and body mass index. Within-pair analyses showed that genetic confounding heterogeneously affected these associations, with six multi-omic factors remaining significantly associated with EAA independent of genetic effects. Replication in older twins showed that some of these associations persist across adult generations.

## Introduction

Globally, life expectancy has increased by one year every three years since the beginning of the 21st century according to the World Health Organization (World Health Organization, 2019). As a result, the number of older people is increasing and is expected to continue to increase in the coming years. Since age is a major risk factor for a variety of diseases (Kaeberlein, 2017), such as COVID-19 and Alzheimer’s disease (Williamson et al., 2020; Hou et al., 2019), such a situation represents a pressing public health and societal issue. A better understanding of the biological mechanisms of aging which progresses at different rates for different people is needed. Thus, a better grasp of the role of genetics and the environment in variation of aging, is a key to addressing this challenge.

The advent of high-throughput technologies has made it possible to generate large amounts of so-called omics data, which ultimately makes it possible, for example, to assess the effect of genes on health using genotype data. Unlike the genotype, which remains essentially the same from birth to death, methylation of the DNA, a common epigenetic modification, has been shown to be dynamic across life span. Both genotype and environmental factors (e.g., diet or early life experiences) interact to shape DNA methylation patterns during life course, giving them a high potential to be used as biomarkers of environmental exposures, behavior or disease. For example, DNA methylation patterns have been identified as biomarkers for smoking and perfluoroalkyl substances (Tsai et al., 2018; Liu et al., 2022). In addition, many studies have provided evidence for age-related changes in DNA methylation levels at specific CpG sites (Christensen et al., 2009). This led to the development of DNA methylation-based epigenetic clocks with the goal of estimating biological age, which is a more accurate reflection of health and a better biologically informed measure of age than chronological age (Salameh et al., 2020). Because the study of biological age can be insightful in comparison to chronological age, another measure of the rate (or pace) of aging is commonly used and referred to as epigenetic age acceleration (EAA), which is determined by regressing biological age on chronological age. Thus, a positive EAA value indicates that an individual is aging faster than expected based on their chronological age.

Several algorithms have been developed to estimate biological aging from DNA methylation data, including six that we used in the current study: Horvath (Horvath, 2013), Hannum (Hannum et al., 2013), GrimAge (Lu et al., 2019), GrimAge2 (Lu et al., 2022), PhenoAge (Levine et al., 2018) and DunedinPACE (Belsky et al., 2022). The EAA estimates derived from each of these clocks have been widely used in studies targeting on phenotypes such as lifestyle (Drouard et al., 2023a), environmental exposures (de Prado-Bert et al., 2021) and diseases (Lundgren et al., 2022; Yusupov et al., 2023; Joyce et al., 2021; Foster et al., 2023; Carbonneau et al., 2024), all of which have demonstrated adverse health outcomes in individuals with greater EAA. However, studies investigating associations between EAA and multiple omics data remain scarce. The identification of omic biomarkers for DNA methylation-based aging would provide a more holistic picture of the biological determinants of aging and a deeper understanding of the underlying biological processes that lead to acceleration of biological age. It would also improve our ability to inform public health policies to prevent accelerated aging of populations, and to develop drug-targeted interventions to reduce the rate of biological aging in patient groups.

Mavromatis and colleagues (Mavromatis et al., 2023) investigated associations of EAA with genomic, transcriptomic and metabolomic data and reported several genes and metabolites to be associated with EAA and longevity. However, mendelian randomization analysis did not indicate any causal relationships between metabolites and EAA. Whether metabolites and plasma biomolecules are causally associated with EAA is therefore largely undetermined. Biological age has also been shown to be heritable (McCartney et al., 20-21; Marioni et al., 2015), but the influence of genetic and environmental factors on the associations between EAA and several layers of the multi-ome remains underexplored. In addition, whether genetics and environment influence these associations similarly across the different omic layers, and whether these associations are consistent across different EAA estimates and human generations are to be investigated.

We designed a multi-omic study of EAA in a twin cohort (Fig. 1). First, we sought to identify markers of EAA in young adults (mean age: 22.3 years old) in four domains: the plasma proteome, the plasma metabolome, the external physical and social exposome, and lifestyle-related traits. We quantified associations between the multi-ome and six different estimates of EAA using linear mixed-effects models. We then investigated the role of genetics and the environment in the observed associations using twin study designs. Finally, we used an independent cohort of older (mean age: 62.3 years old) twins with available EAA, proteomic, and metabolomic data with the aim to replicate the associations identified in the young twins. As the age difference between the two twin cohorts is substantial, this allowed us to discuss which markers of EAA were consistent across generations and whether environmental and genetic influences on the associations remained the same.

**Fig. 1:**
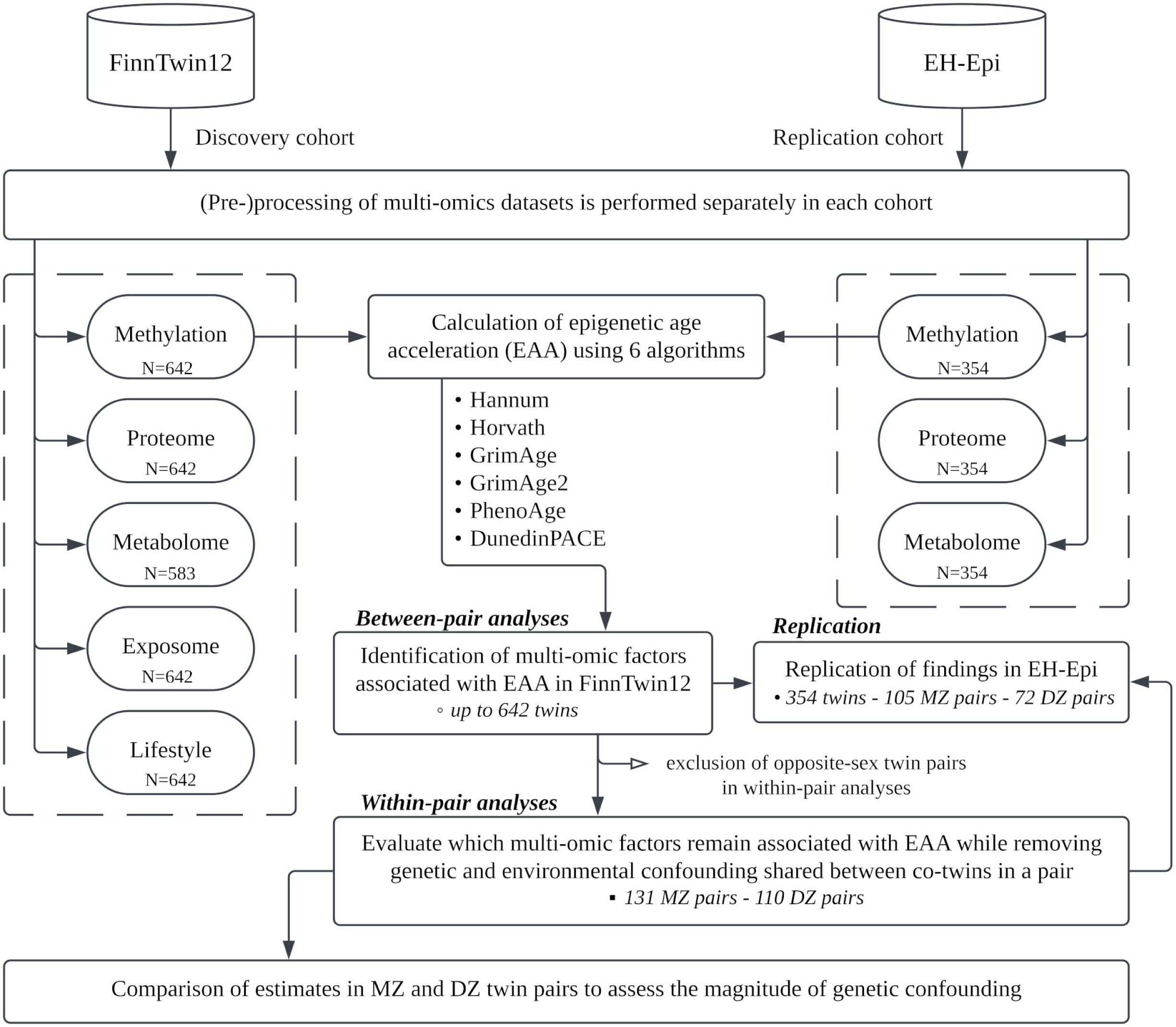
Study flow chart. **Caption:** The study was divided into three stages. The first was to quantify the associations between EAA and multi-omic factors with all twin individuals from the FinnTwin12 cohort, referred to as between-pair analyses. Next, within-pair analyses were carried out on all complete same-sex twin pairs. Finally, replication of the proteins and metabolites identified in FinnTwin12 was performed in the external EH-Epi sample. EAA: Epigenetic Age Acceleration. MZ pairs: monozygotic pairs. DZ pairs: dizygotic pairs.

## Results

The main analyses were divided into three parts (Fig.1) and involved two independent samples of twins, as described in Table 1. First, associations between EAA and multi-omic factors were quantified in all twin individuals from FinnTwin12. These analyses are referred to as between-pair analyses. A list of the multi-omic factors included in these analyses is available in the Supplementary Material (Table S1). Second, to investigate whether associations were robust to correction for shared environmental and genetic effects, within-pair analyses were performed. In parallel, we investigated the contribution of genetics in the observed associations by comparing within-pair estimates in monozygotic (MZ) twin pairs to those in dizygotic (DZ) twins pairs, as MZ twins in a pair are identical at the DNA sequence level, whereas DZ twins in a pair share on average only half of their segregating genes. Finally, significant associations observed between EAA and proteins and metabolites were examined in a replication set of older twins from the Older Cohort EH-Epi sample.

**Table 1:** Description of the FinnTwin12 and EH-Epi samples. **Caption:** Variable descriptions are given in terms of mean and standard deviation (sd) for continuous variables, and in terms of frequencies for binary variables marked with the symbol *.

|  | **FinnTwin12 (N=642)** | **EH-Epi (N=354)** |
| --- | --- | --- |
|  | mean (sd) / Frequencies | mean (sd) / Frequencies |
| Age at blood sampling | 22.3 (0.6) | 62.3 (3.8) |
| Body mass index | 23.2 (3.9) | 27 (4.9) |
| Current smokers * | 249 | 56 |
| Never smokers * | 102 | 168 |
| Sex (females) * | 386 | 212 |
| Complete MZ pairs * | 131 | 105 |
| Complete DZ pairs * | 110 | 72 |

### Between-pair analyses reveal a broad range of multi-omic associations with EAA

Linear mixed-effects models were fitted to quantify associations between six different EAA estimates (Fig. 1) and the multi-ome in the FinnTwin12 participants. Models were adjusted for sex, body mass index (BMI) and smoking. We identified 40 unique multi-omic factors associated with one or multiple EAA estimates after correcting p-values for multiple testing (total number of associations: 77) (Fig. 2, Table S2). All omics showed associations with at least one EAA measure, with the DunedinPACE estimate of EAA showing the greatest number (n=19) of associations with multi-omic factors (Table S2). As could be expected, the clocks that were developed to estimate chronological age (Hannum and Horvath), showed significantly fewer associations compared with the clocks that were developed to estimate mortality and phenotypic aging (GrimAge, GrimAge2 and PhenoAge) (Table S2). Similarities and differences between the EAA estimates in their associations with multi-omic factors are shown in Fig. 3. The direction of the observed multi-omic factor associations shared across the EAA estimates were consistent (i.e., the sign of the coefficients was the same across all the EAA estimates, Table S2).

**Fig. 2:**
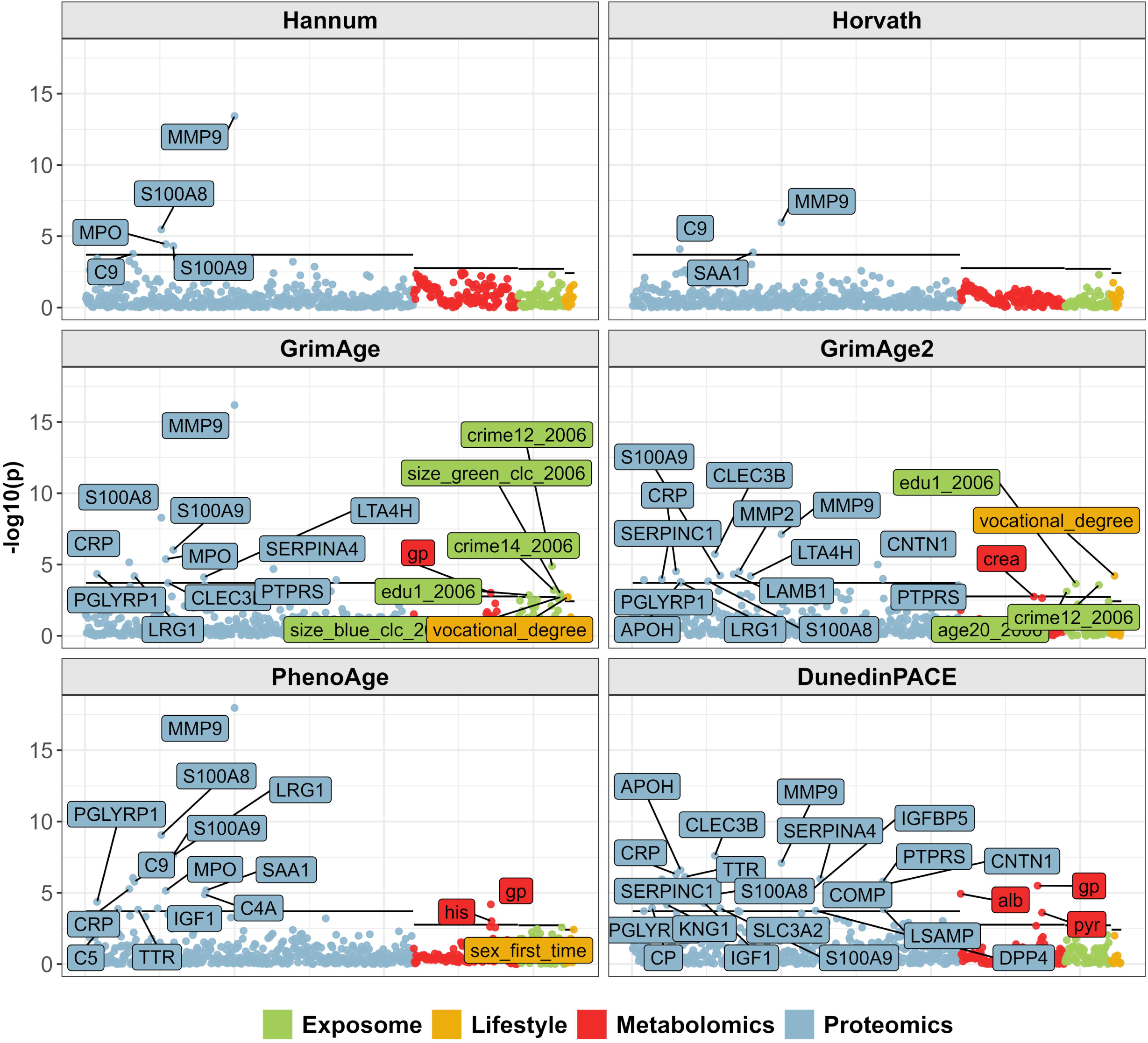
Between-pair analyses reveal numerous associations between epigenetic age acceleration and multi-omic factors. **Caption:** Linear mixed-effect models were used to quantify associations between EAA and multi-omics factors in between-pair analyses. Sex, body mass index and smoking were used as covariates. Correction for multiple testing was based on the number of principal components needed to cover 95% of the initial variance for each omic, and is indicated by a solid line. A description of the multi-omics factors is available in the supplementary material (Table S1). EAA: epigenetic age acceleration. p: p-value resulting from testing the nullity of the multi-omic factor coefficient.

**Fig. 3:**
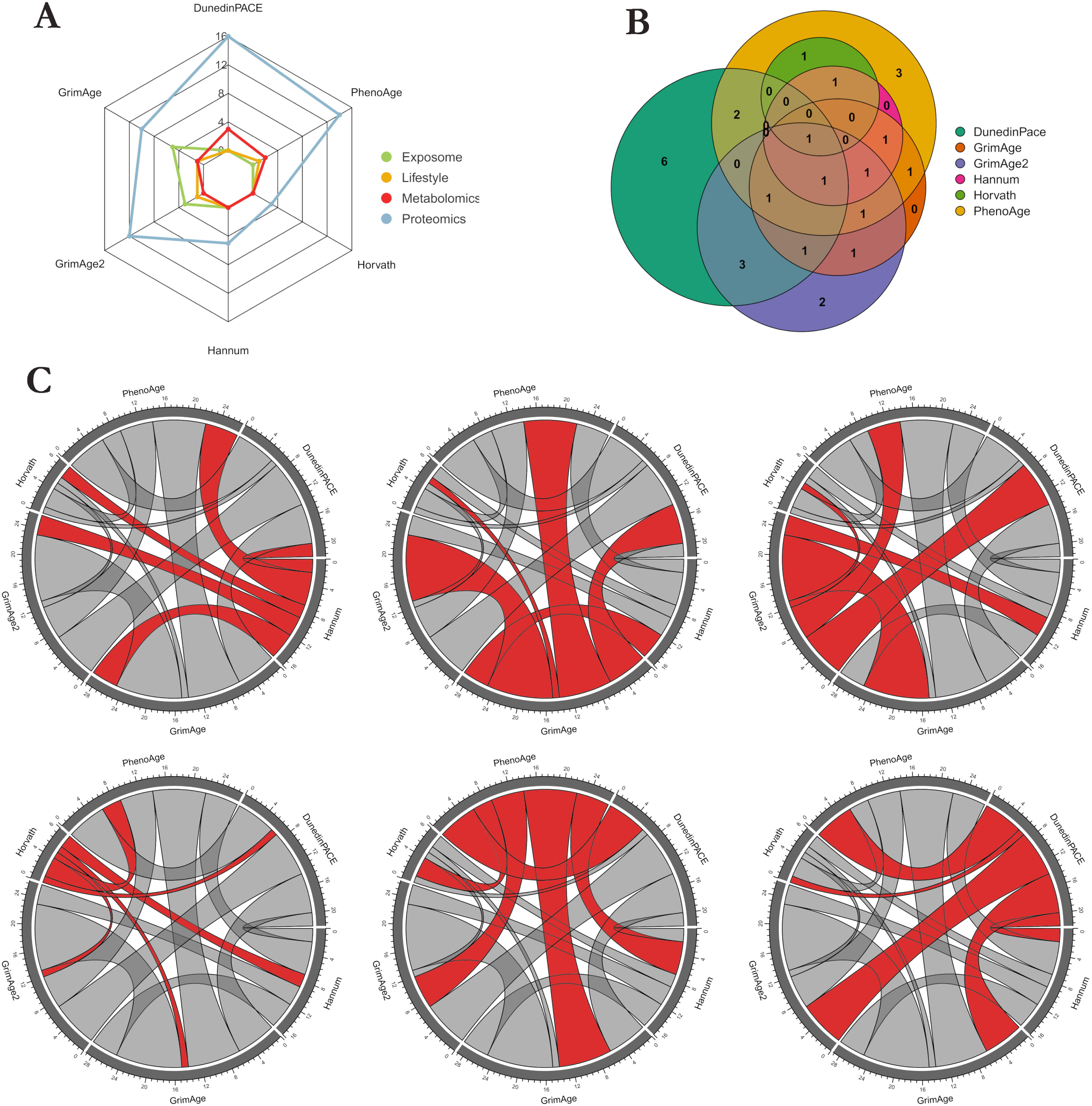
Common associations between multi-omic factors and EAA across different EAA estimates, and omic specificities of different EAA estimates. **Caption: (A)** Radar plot showing the number of associations between EAA and proteins, metabolites, and lifestyle or exposome variables for each EAA estimate. Only the GrimAge and GrimAge2 estimates were associated with exposome variables, whereas the DunedinPACE estimate showed a greater number of associations with plasma omics. **(B)** Euler plot of the overlapping identified proteins across the 6 EAA estimates. 62% of these proteins were associated with more than one EAA estimate. **(C)** Circos plots showing the pairwise number of shared associations between EAA estimates.The six circos plots are identical, with connections between an EAA estimate and others colored in red. The thicker the connection between two EAA estimates, the more they share common associations with multi-omic factors. For example, the DunedinPACE EAA estimate is associated with several multi-omic factors that are also associated with the GrimAge, GrimAge2, and PhenoAge estimates, but very few with Hannum and Horvath (see circos plot at the bottom right).

### Associations between proteins and EAA estimates

Altogether 60 associations were identified between EAA estimates and plasma proteins. These involved 28 unique proteins, of which 11 were associated with only one estimate of EAA. The rest were found to be associated with multiple EAA estimates (Fig.3B). Here, the protein nomenclature will follow the gene nomenclature for clarity. Matrix metallopeptidase 9 (coding gene: *MMP9*, with the respective protein referred to as MMP9) was the only protein associated with all six EAA estimates, with standardized coefficients ranging from 4.9 to 9.1. Proteins associated with three or more EAA estimates also included S100-A8 (S100A8), S100-A9 (S100A9), C-reactive protein (CRP), complement component C9 (C9), leucine-rich alpha-2-glycoprotein (LRG1), myeloperoxidase (MPO), and receptor-type tyrosine protein phosphatase S (PTPRS). The full set of proteins associated with EAA estimates and detailed summary statistics are available in the supplementary material (Table S2).

### Associations between metabolites and EAA estimates

Significantly fewer associations were observed between metabolites and EAA estimates compared with proteins and EAA estimates, and none of the associations were with EAA estimates developed for chronological age. Four metabolites, albumin (variable: alb), glycoprotein acetyls (gp), histidine (his) and pyruvate (pyr) were associated with EAA estimates (Fig. 2 and Table S2). Glycoprotein acetyls was the only metabolite associated with multiple EAA estimates (standardized coefficient range: 3.2-4.4): PhenoAge, GrimAge and DunedinPACE (Fig. 3).

### Associations between exposome and lifestyle, and EAA

Several multi-omic factors associated with EAA other than proteins and metabolites were also identified (Fig.2; Table S2). Associations between the external exposome and EAA were identified only with GrimAge and GrimAge2 estimates. Neighborhood crime rates per capita at the municipal level (crimes against life and health: crime12_2006; crimes against authority and public order: crime14_2006), proportion of individuals aged 20 to 29 (age20_2006) and proportion of the population with unknown or no education after primary or lower secondary education in the population aged 16 and over (edu1) at the postal code level, and areas of the nearest green (size_green_clc_2006) and blue (size_blue_clc_2006) spaces were associated with EAA. Associations with EAA were positive for edu1 and size_green_clc_2006, otherwise negative. Finally, we identified two lifestyle-related variables associated with EAA. Twins who had their first sexual intercourse before the age of 18 had a higher PhenoAge EAA than those who had their first sexual intercourse after the age of 18 (standardized coefficient: 2.9). Twins who had a vocational degree were also found to age faster than twins who did not have a vocational degree (standardized coefficient with GrimAge: 3.1; with GrimAge2: 4.0).

### Effects of BMI and smoking

We also performed additional between-pair analyses adjusting only for sex to assess the confounding effects of BMI and smoking on the association between EAA and the multi-ome. These analyses resulted in the identification of 142 unique multi-omic factors associated with EAA, including 89 proteins, 41 metabolites, 7 exposome variables and 5 lifestyle-related variables (Table S3). While the increase in the number of exposome and lifestyle variables was modest compared to the fully adjusted models, not adjusting for BMI and smoking dramatically increased the number of associations with plasma omics. These results suggest that most of the associations between the plasma omics and EAA in the models adjusted for sex only are likely to reflect the effects of BMI and smoking habits on EAA, the proteome and the metabolome.

### Within-pair models reveal associations robust to correction for shared environmental factors and genetics

We used within-pair analyses to investigate whether there would be significant associations between EAA and the multi-ome while correcting for shared environment and genetics. We performed three sets of within-pair analyses with different degrees of correction for shared environmental and genetic effects: all same-sex twin pairs, same-sex DZ twin pairs only, and MZ twin pairs only. While within-pair analyses using all pairs correct for environmental factors and genetic polymorphisms that are shared between the co-twins while maintaining a high level of statistical power, only within-pair analyses using MZ pairs additionally ensure the removal of all genetic confounding in the associations.

A total of 53 associations were significant in the within-pair analyses using all pairs of twins (n= 241 pairs), involving 31 multi-omic factors (Table S4). About half of these associations (27/53) were also identified in between-pair analyses (Fig.S1). These findings suggest that environmental factors and genetic polymorphisms shared by the co-twins either do not have an influence, or cannot be the only influence, on part of the associations between EAA and the multi-ome. Significant associations found in both between-pair and within-pair analyses were mostly between proteins and EAA estimates, with the exception of the metabolite albumin which was negatively associated with DunedinPACE estimate of EAA. MMP9 was associated with all six EAA estimates, as observed in the between-pair analyses.

In the within-pair analyses of DZ pairs only (n=110 pairs), 23 associations were significant (involving 14 multi-omic factors), 16 of which were with proteins (Table S5). We observed associations between several crime rate variables from the exposome domain and EAA, as well as an association between the lifestyle-related variable indicating participation in clubs and Horvath estimate of EAA, all of which were negative.

In MZ pairs only (n=131 pairs), we observed 8 significant associations (involving six multi-omic factors), three of which were with proteins, three with metabolites, and two with exposome variables (Table S6). MMP9 was positively associated with GrimAge (standardized coefficient: 5.5) and PhenoAge (standardized coefficient: 5.7) estimates of EAA. The complement component C6 (C6) protein was positively associated with GrimAge’s estimate of EAA (standardized coefficient: 4.0). Regarding metabolites: histidine was negatively associated with PhenoAge’s estimate of EAA (standardized coefficient: –3.3). Glycoprotein acetyls metabolite was negatively associated with DunedinPACE’s estimate of EAA (standardized coefficient: –3.3), and lactate was positively associated with Phenoage’s estimate of EAA (standardized coefficient: 3.3). These results suggest either that plasma factors, such as MMP9 and histidine, are causally related to EAA, or that environmental factors not shared between the co-twins influence these associations. Finally, we identified that the proportion of individuals aged 90 and older in the area linked to the twins residential geocode (exposome variable age90_2006), was positively associated with both PhenoAge (standardized coefficient: 3.3) and GrimAge (standardized coefficient: 3.4) estimates of EAA within MZ pairs. This echoes the negative associations that were identified in between-pair analyses between the proportion of young individuals aged 20 to 29 in the geocode (age20_2006) and EAA.

### Within-pair analyses provide evidence for heterogeneous genetic influences in the observed associations

While within-pair analyses allow for the identification of associations between EAA and the multi-ome that are independent of shared environmental and genetic factors, they also provide insight into how genetics underlie these associations: larger estimates in DZ pairs compared with MZ pairs reflect greater genetic influence in the associations. Therefore, we examined in detail the associations that were significant in all twin pairs and compared the estimates of these associations in MZ twin pairs only with those in DZ twin pairs only (Fig.4A, Fig.4B). Of the 53 significant associations identified in within-pair analyses in all twin pairs, 49 showed higher estimates in DZ pairs compared with MZ pairs (Table S7). On average, the estimates in DZ pairs were 76% higher than in MZ pairs (interquartile range (%): 17.5-87.7).

**Fig. 4:**
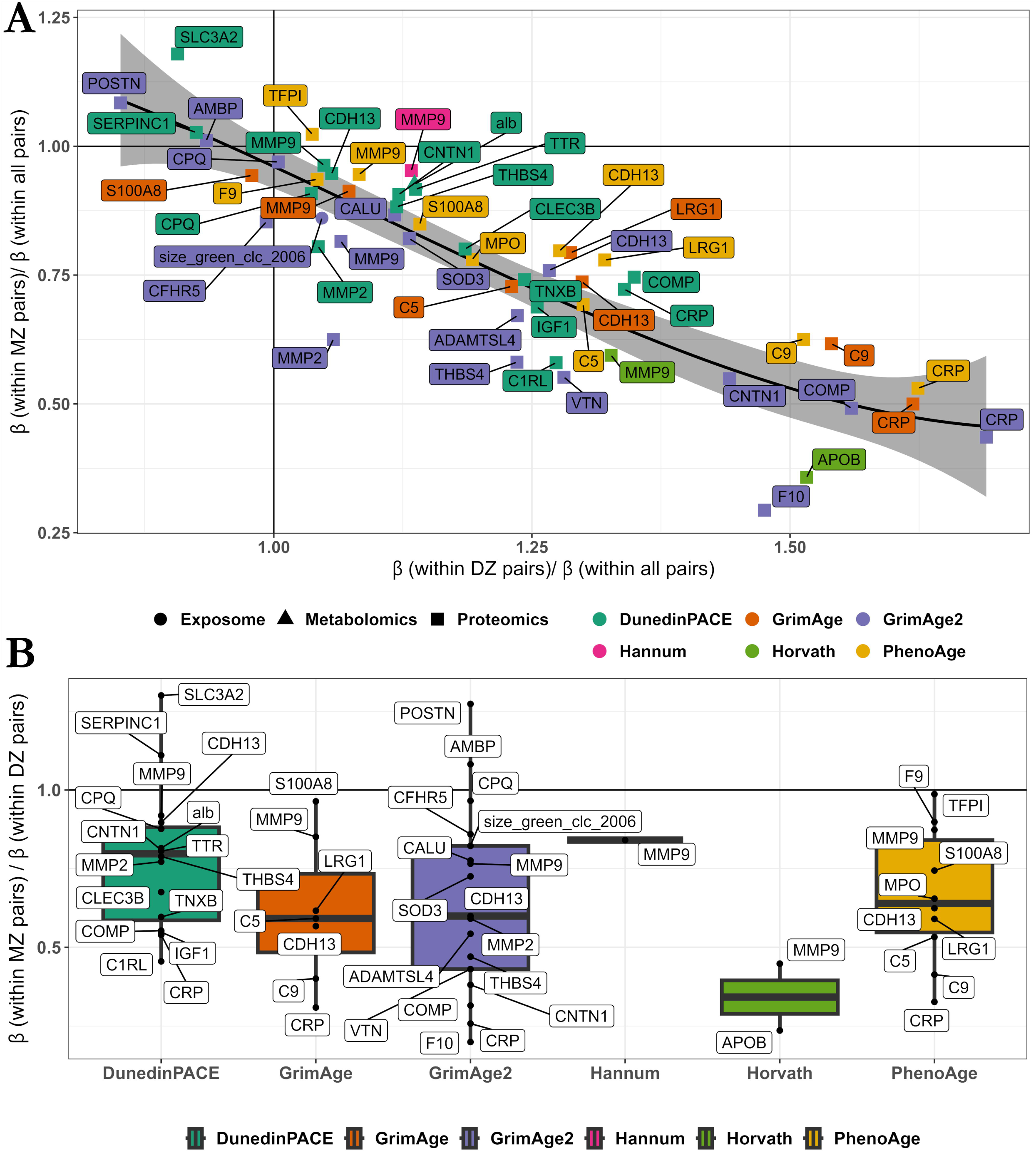
Comparisons between MZ and DZ pairs in within-pair analyses provide evidence for genetic confounding in associations. **Caption: (A)** Comparison of estimates in MZ [β(within MZ pairs)] and DZ [β(within DZ pairs)] pairs for multi-omics factors significantly associated with EAA in all pairs [β(within all pairs)]. Multi-omics factors that are located the farthest down and to the right are those for which the genetic effects in the association are the most intense. Conversely, weak genetic effects in associations are indicated by multi-omics factors with coefficient ratios close to 1. Black line indicates a fitted three-degree polynomial curve. **(B)** For most multi-omic factors associated with EAA in all pairs, estimates in DZ twins [β(within DZ pairs)] are higher than those in MZ twins [β(within MZ pairs)]. The lower the ratio of MZ to DZ pair coefficients, the more likely the genetic confounding in these associations with EAA. EAA: epigenetic age acceleration. Variable descriptions are available in the supplementary material (Table S1).

Thirteen associations, involving 10 unique proteins, had estimates in DZ pairs that were at least twice as large as in MZ pairs (Fig.4B, Table S7), indicating that genetic effects are likely the main and only driver for these associations. These proteins were CRP, coagulation factor X, cartilage oligomeric matrix protein (COMP), apolipoprotein B-100 (APOB), C9, vitronectin, thrombospondin-4, CNTN1, complement C1r subcomponent-like protein, and MMP9. Interestingly, while the association between MMP9 and Horvath EAA showed a high degree of genetic confounding (ratio coefficient in MZ pairs vs. DZ pairs: 0.45; Fig.4B), the associations of MMP9 with other clocks showed a lower degree of genetic influence (ratio coefficient in MZ pairs vs. DZ pairs ranged from 0.77-0.92 for the remaining five clocks; Fig.4B). This suggests that there may be differences in genetic influences between MMP9 and EAA across clocks.

### Replication of associations with plasma proteins and metabolites in an independent sample of older twins

We sought to replicate the findings from the FinnTwin12 study of young adult twins in an independent cohort of older twins (Fig.1; Table 1).

Of the 40 multi-omic factors identified in the between-pair analyses in FinnTwin12, 4 were metabolites and 28 were proteins. Of these, all 4 metabolites and 15 proteins could be investigated in the EH-Epi sample (Fig.5A) due to proteomics platform differences between the cohorts. The standardized coefficients estimated in this independent EH-Epi twin sample correlated highly with those from FinnTwin12 (Pearson correlation estimate: r=0.73; confidence interval: [0.55,0.85]). Altogether, 17 of the 39 associations replicated in the EH-Epi sample with nominal p-value below 0.05 (Fig.5A, Table S8). Nine associations were robust to Bonferroni correction, six of which were with proteins. MMP9 was positively associated with DunedinPACE (standardized coefficient: 3.8; Bonferroni p=0.006) and GrimAge2 (standardized coefficient: 3.6; Bonferroni p=0.013) estimates of EAA. CNTN1 was negatively associated with DunedinPACE (standardized coefficient: –3.6; Bonferroni p=0.014) and GrimAge2 (standardized coefficient: –3.9; Bonferroni p=0.004) estimates of EAA. PTPRS was associated with both GrimAge (standardized coefficient: –4.0; Bonferroni p=0.003) and GrimAge2 (standardized coefficient: –3.7; Bonferroni p=0.010) estimates of EAA. Among metabolites, the glycoprotein acetyls metabolite was associated with both GrimAge (standardized coefficient: 4.6; Bonferroni p=2.2e-4) and DunedinPACE (standardized coefficient: 4.5; Bonferroni p=3.7e-4) EAA. In addition, albumin was negatively associated with DunedinPACE’s EAA (standardized coefficient:-4.6; Bonferroni p=2.7e-4).

**Fig. 5:**
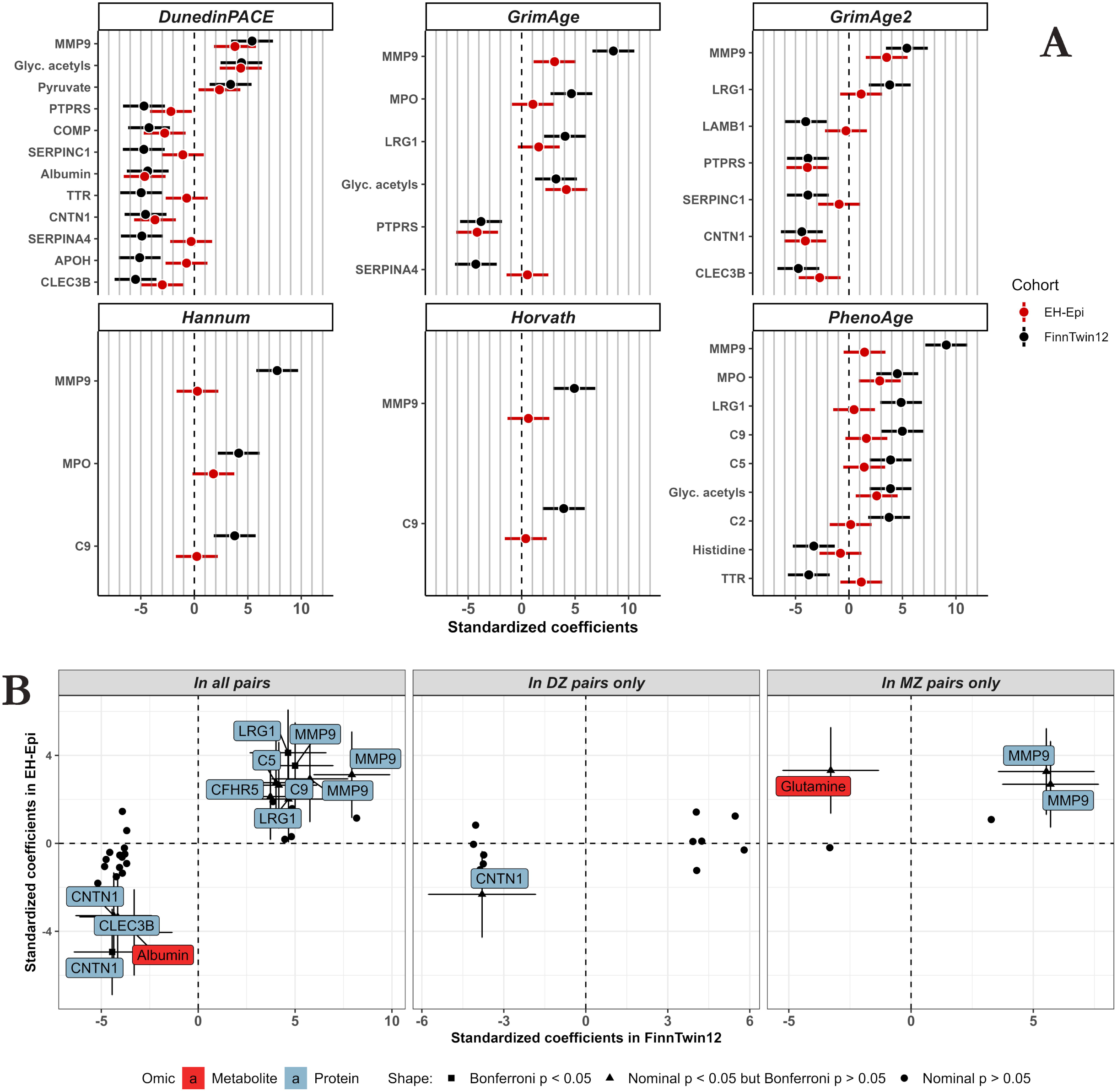
Replication of between-pair and within-pair findings from FinnTwin12 young adult twins in the EH-Epi sample of older adults. **Caption: (A)** Standardized coefficients and their 95% confidence intervals in the EH-Epi sample next to those found in FinnTwin12. Almost half of the replicated associations had a nominal p-value testing the null of the coefficients below 0.05, 9 of which were robust to Bonferroni correction. **(B)** Comparison of standardized coefficients obtained in within-pair analyses in the EH-Epi sample with those found in FinnTwin12 in three configurations: in all pairs, in DZ pairs only, and in MZ pairs only. Associations with nominal p-values below 0.05 are labeled and their 95% confidence intervals are shown. The shape of the dots indicates whether the associations were robust to the Bonferroni correction considering all associations tested. Metabolites are annotated with their complete name, with the exception of Glyc. acetyls, being glycoprotein acetyls. Proteins are annotated with their coding genes. *C9*: complement component C9. *MPO*: myeloperoxidase. *MMP9*: Matrix metalloproteinase-9. *LRG1*: Leucine-rich alpha-2-glycoprotein. *CLEC3B*: tetranectin. *SERPINA4*: kallistatin. *PTPRS*: receptor-type tyrosine-protein phosphatase S. *SERPINC1*: antithrombin-III. *APOH*: beta-2-glycoprotein 1. *LAMB1*: laminin subunit beta-1. *CNTN1*: contactin-1. *C5*: complement component C5. *TTR*: transthyretin. *DPP4*: dipeptidyl peptidase 4. *COMP*: cartilage oligomeric matrix protein. *CFHR5*: complement factor H-related protein 5.

In within-pair analyses, we were able to test a total of 53 associations in the EH-Epi sample out of the 84 identified in the FinnTwin12 including all pairs, DZ, and MZ pairs only (Fig.5B, Table S9). Standardized coefficients in the EH-Epi sample were moderately correlated with those obtained in FinnTwin12 (Pearson correlation estimate: r=0.67; confidence interval: [0.49,0.80]). A total of 16 associations were replicated with a nominal p-value below 0.05 (Fig.5B) and four associations were robust to Bonferroni correction (i.e., nominal p-value below 0.05/53= 9.4e-4) in within-pair analyses using all twin pairs. CNTN1 was most strongly associated with EAA (clock: GrimAge2; standardized coefficient: –4.9; Bonferroni p=1.0e-4), followed by LRG1 (GrimAge; standardized coefficient: 4.1; Bonferroni p=0.003), albumin (DunedinPACE; standardized coefficient: –4.1; Bonferroni p=0.004) and MMP9 (DunedinPACE; standardized coefficient: 3.5; Bonferroni p=0.028). In addition, associations within MZ pairs showed high evidence of significance despite not reaching the Bonferroni threshold, such as the metabolite glutamine (nominal p=1.3e-3; Bonferroni p=0.067) and MMP9 (nominal p=1.5e-3; Bonferroni p=0.079) with DunedinPACE and GrimAge EAA estimates, respectively. Interestingly, the association between glutamine and DunedinPACE EAA was negative in the FinnTwin12 sample of young adults, but positive in the older twins of the EH-Epi sample.

## Discussion

Our study provides a comprehensive, in-depth epidemiological view of the connections between EAA and the multi-ome. We identified 40 unique multi-omic factors associated with accelerated epigenetic aging – including proteins, metabolites, external exposures, and lifestyle-related traits – while adjusting for age, BMI, and smoking. A larger number of associations was identified through analyses not corrected for BMI and smoking, suggesting that most of such associations between plasma omics and EAA likely reflect body weight and smoking habits. Using twin study designs, we evaluated the influence of genetic and environmental factors on multi-omic associations with EAA, and demonstrated that these associations show varying degrees of evidence for genetic confounding. In addition, we identified eight significant associations with EAA in within-pair analyses of MZ twins, suggesting either causality in these associations and/or unique environmental factors influencing them independently of genetic factors. Finally, by replicating our findings obtained in a sample of 20-year-old twins in an independent sample of 60-year-old twins, we demonstrated that a small proportion of multi-omic factors reflect accelerated epigenetic aging across age groups.

We identified 28/439 proteins associated with EAA in between-pair analyses. Most of these proteins were associated with more than one estimate of EAA, such as MMP9, which was strongly associated with all six estimates of EAA. This is consistent with the known role of metalloproteinases in aging (Cancemi et al., 2020; Freitas-Rodriguez et al., 2017), but also echoes for example how the GrimAge and GrimAge2 clocks were developed (Lu et al., 2019; Lu et al., 2022), since they involved the use of tissue inhibitors of metalloproteinases, which are regulators of metalloproteinases. Associations between MMP9 and EAA estimates were present in within-pair models as well, two of which (with PhenoAge and GrimAge) remained significant within MZ pairs. This suggests that the associations of MMP9 with PhenoAge and Grimage estimates of EAA are either causal or at least partially due to environmental effects not shared between the siblings. Attempts to replicate these two associations in older MZ twin pairs of the EH-Epi sample failed to reach Bonferroni significance despite nominal p-values below 0.05 (GrimAge: nominal p=1.5e-3, Bonferroni p=0.08; PhenoAge: nominal p=8.4e-3, Bonferroni p=0.45) and effect sizes in the same direction (i.e., positive) as in FinnTwin12.

PTPRS associated negatively with three EAA estimates (GrimAge, GrimAge2 and DunedinPACE) in the FinnTwin12 sample, and the finding was replicated in the EH-Epi for GrimAge and GrimAge2. PTPRS plays a multifaceted role in aging through its impact in neural development and regeneration (Shen et al., 2009; Martin et al., 2011), metabolic regulation and inflammation (Zheng et al., 2024). Its dysregulation can lead to inflammaging, changes in metabolism and age-related diseases. LRG1 was positively associated with three EAA estimates (GrimAge, GrimAge2 and PhenoAge) in FinnTwin12, and associations with both GrimAge and PhenoAge remained significant in the within-pair analysis of all FinnTwin12 pairs and with GrimAge in all EH-Epi pairs. LRG1 is an important factor of innate immunity, where it has been postulated to act as an acute phase protein, responding to infections and other inflammatory stimuli (Codina et al., 2010). It has been further shown to be upregulated in a plethora of human diseases and to contribute to the disease process in multiple organs (Camilli et al., 2022). CNTN1 showed consistent negative association with DunedinPACE and GrimAge2 in both FinnTwin12 and EH-Epi, and it was replicated in the within-pair analyses. CNTN1 has been previously associated with both frailty (Liu et al., 2024) and frailty trajectories (Varghese et al., 2021) in proteomic studies in mid– and late-life. These and other studies (Osawa et al., 2020) provide evidence for CNTN1 having a protective role in the development of frailty, which has been associated with accelerated EAA (Gale et al., 2018). These findings are in line with our observations on higher plasma CNTN1 levels associated with slower EAA both in mid-life and late-life.

A total of 4/140 metabolites were associated with EAA in between-pair analyses. Glycoprotein acetyls were positively associated with EAA in both early and late adulthood in our study. Glycoprotein acetyls are markers of inflammation and they may contribute to inflammaging, the low grade inflammation occurring during aging predisposing to many age-related diseases (Franceschi et al., 2014). A very recent study demonstrated that glycoprotein acetyls are positively associated with frailty index in older age (Bålsrud et al., 2024), however, no study has associated glycoprotein acetyls with EAA before. While our findings are supported by the previous studies on inflammaging and frailty in older age, it also adds to the literature by demonstrating a direct link between glycoprotein acetyls and EAA, which is present already in early adulthood.

We observed both in the younger and older twins that the higher the albumin levels, the lower the EAA, which is consistent with the literature on epigenetic aging (Uchehara et al., 2023) and aging in general (Lau et al., 2024), where lower albumin levels are associated with higher calendar age. The same has been observed for mortality risk among healthy adults at all ages (Fulks et al., 2010). We verified the negative association between albumin and EAA in within-pair analyses in all younger and older twin pairs, but no significant differences were observed within MZ twin pairs, and therefore genetic confounding could not be completely ruled out in these associations.

Pyruvate was positively and histidine negatively associated with EAA in the current study. To our knowledge, these metabolites have not been previously reported to associate with EAA. However, histidine has been shown to predict chronological age (Lau et al., 2024). In the current study histidine was significantly associated with the PhenoAge estimate of EAA in both between-pair and within-pair analyses of MZ twins suggesting direct effect between histidine level and EAA, independent of genetic effects. This may be in contrast with the findings of a multi-omic study of EAA that did not identify causal associations between any metabolite and EAA using Mendelian randomization (Mavromatis et al., 2023). Both study designs can, however, only provide theoretical proof for causation, which could explain this discrepancy.

Another metabolite we identified to differ within the MZ twin pairs was glutamine, which was negatively associated with the DunedinPACE estimate of EAA in FinnTwin12. In contrast, when we attempted to replicate this association in older twins of the EH-Epi sample, we found a substantial (but not significant after Bonferroni correction) positive association. A positive association between glutamine with chronological age was also observed in a large metabolomic study of aging including adults ranging 24-86 years olds (Lau et al., 2024). This suggests that the role of glutamine in biological aging may change over decades, with higher levels indicating a lower rate of aging in young adulthood but a higher rate of aging in older adults. Further studies to disentangle the relationship between glutamine and biological aging are needed, particularly in younger populations.

Lifestyle variables were also associated with EAA. We observed a greater acceleration of epigenetic aging (PhenoAge) in individuals who had their first sexual intercourse before the age of 18 in between-pair analyses. This echoes the results of a study based on women from the ALSPAC cohort, in which an earlier age at first sexual intercourse was associated with greater EAA during childhood (Schlomer, 2024). Another study showed the same direction of association in women and demonstrated the role of BMI as a mediator in this association (Zhang et al., 2023). Thus, our findings consolidate the existing literature by showing that earlier age at sexual intercourse is associated with greater EAA in a sample including both women and men, and adjusting for BMI but also for smoking. The other lifestyle variable that we observed to be significantly associated with EAA was the variable representing holding a vocational degree, which was positively associated with greater EAA calculated with GrimAge and GrimAge2. This is consistent with the known association between lower education and greater GrimAge EAA reported in the literature (Crimmins et al., 2021). Associations between EAA and lifestyle variables, such as education level and age at first sexual intercourse, may collectively reflect an underlying association between lower socioeconomic status and higher biological aging (Schmitz et al., 2022).

We identified multiple associations between the external exposome and the GrimAge and GrimAge2 estimates of EAA in the between-pair analyses. Significant associations were observed with crime rates, the proportion of 20-year-olds, sizes of nearest blue and green spaces areas, and average educational attainment, all in the residential neighborhood indexed by geocode. For all these or related characteristics, similar associations have been reported (Kim et al., 2023; Dos Santos Oliveira et al., 2023; Jovanovic et al., 2017). For instance, we observed that the higher the proportion of individuals with no education after primary or secondary education among individuals over 16-years old at the geocode level, the higher the acceleration of epigenetic aging of individuals living in such an area. Our study therefore suggests that lower educational attainment both at the geocode level (Exposome domain) and at the individual level (Lifestyle domain) is associated with greater EAA.

Within-pair analyses showed only a few associations between EAA and the external exposome, suggesting that a large proportion of the associations observed in the between-pair analyses are likely due to shared environmental effects and genetic confounding. However, the proportion of individuals aged over 90 years at the geocode level was significantly associated with increased EAA in within-pair analyses in MZ twin pairs, suggesting a strong association between age structure in a geocode and EAA independent of all genetic and shared environmental factors. While the literature on EAA in relation to the prenatal, childhood or adolescent exposome is growing, that is not the case for the adult exposome. Further studies between EAA and the adult external exposome are therefore warranted.

While some multi-omic associations are shared between different EAA estimates, we also observed differences at the level of domains of the multi-ome. As each EAA estimate is constructed using different populations and outcomes, they characterize different aspects and functions of biological aging, resulting in greater affinity to either deep (i.e., molecular) or shallow (e.g., external exposome) omics. Hannum and Horvath clocks that were trained to predict chronological age only captured age acceleration –related alterations in proteins. All the observed associations attenuated in the within-pair analyses of MZ twin pairs. This may reflect rather strong effects of underlying genetics, which affect methylation at age-associated CpG sites as well as protein expression. Genetic effects may be more direct and less affected by the environment for clocks that estimate calendar age compared with the clocks that were trained to predict phenotypes that reflect the effect of aging on health and mortality risk. Also, the DunedinPACE estimate of EAA showed the strongest connections with metabolites and proteins. Conversely, The GrimAge, GrimAge2 and PhenoAge estimates of EAA were the only clocks whose connections with the external exposome and lifestyle-related traits were significant. Our study therefore provides an excellent platform for assessing which omics of the multi-ome, shallow or deep, and which EAA clock is best suited to a given research question. Extending multi-omic approaches to epigenetic aging to other omics than those investigated in the current study may lead to a better understanding of biological aging, and such attempts should be fostered.

Our study had limitations that may have affected its results. The first is sample size, as the number of twin pairs included in the analyses was modest with regards to the number of variables included in our analyses, and therefore to the number of tests performed. However, twin cohorts with metabolomic, proteomic, external exposome, and lifestyle data in a similar or larger number of twins are rare, demonstrating the uniqueness of our study and its unprecedented contribution to the literature. In addition, since the FinnTwin12 twins are young adults, they have had only limited time to build their own experiences outside of their familial environment which the twins in a pair share. This may have led to relatively modest within-pair differences in the omic and EAA profiles, and therefore a failure to detect significant associations. Another limitation that prevented us from validating all the observed associations in the FinnTwin12 sample in the independent sample (EH-Epi) is the fact that the same exposome and lifestyle data were not available for both samples, and the proteomic data were generated using different platforms.

In conclusion, our study provides an in-depth view of the connections that link the multi-ome and acceleration of epigenetic aging. We have shown that associations between multi-omic factors and EAA are due to different contributions of genetic and environmental factors, which may ultimately allow the identification of biomarkers of interest, e.g. when conducting intervention or genetic studies. However, whether the determinants of biological aging in young adults are similar to that in children or in old adults remains to be further explored. Although the replication of our findings in an independent sample of 60-year-old participants partially met this challenge, intergenerational studies are encouraged to harmonize findings across age groups.

## Methods

### Cohorts and participants

Main analyses were performed on young adult twins (mean age: 22.3; range: 21.0-24.7) from the FinnTwin12 cohort, including both between-pair and within-pair analyses. Significant associations between EAA and plasma biomolecules (i.e., proteins and metabolites) were replicated in an independent sample of older twins (mean age: 62.3; range: 56.0-70.0) from the Older Cohort EH-Epi sample. Data processing is presented separately for each cohort, but the statistical analysis methodology applies to both cohorts.

### FinnTwin12

FinnTwin12 is one of the core sub-cohorts of the nationwide Finnish Twin Cohort (FTC), whose initial aim was to study adolescent behavior and mental health (Kaprio, 2006; Rose et al., 2019). Participants were first identified from the Digital and Population Data Services Agency of Finland, and later asked to complete a series of questionnaires at the ages of 11/12, 14, 17, and as young adults (mean age: 22 years). In the latest assessment wave, a sample of these twins participated in a more detailed in-person study with clinical measures, interviews, and questionnaires (Rose et al, 2019). In the morning of the assessment, a fasting venous blood sample was drawn, processed, and immediately stored at –80C. Multiple omics were generated, including metabolomic, proteomic, and epigenetic data from plasma and DNA.

The present study is based on 642 twins from FinnTwin12 with all of the aforementioned complete omics data, except for 9% of them with no metabolomics data (Fig.1). All molecular omics were derived from the same blood sample. Other omics data (i.e., lifestyle and external exposome data) were generated to be chronologically consistent with the time at which blood sampling was performed. The sample included 60% female participants. Age at blood sampling ranged from 21.0 to 24.7 years (mean: 22.3) in this sample (Table 1).

### Essential Hypertension Epigenetics (EH-Epi) study

The Old Cohort is another sub-cohort of the FTC, which was initiated in the 70s and for which longitudinal data have been collected over time (Kaprio et al., 2019). Based on the fourth survey in 2011, a sample of this cohort, referred to as EH-Epi, was constructed a few years later to study pairs of twins with a difference in blood pressure, as described elsewhere (Kaprio et al., 2019). These twins were invited to come in for a one-day in-person study, including measurement of their blood pressure. During their visit, the twins completed interviews and questionnaires, and their weight and height were measured. Fasting venous blood samples were also collected, from which multiple omics were generated (Drouard et al., 2022).

We used 379 twins from EH-Epi that had complete DNA methylation, proteomic, and metabolomic data available to replicate some of the findings from the main analyses (Fig.1). The twins in this sample were approximately 40 years older than those in FinnTwin12 (mean: 62.3; range: 56.0-70.0). BMI ranged from 18.1 to 45.9 kg.m^-2^ (mean: 27.0; SD: 4.9) (Table 1).

## Data processing in FinnTwin12

### DNA methylation and EAA calculation

DNA methylation levels were quantified using Infinium Illumina HumanMethylation450K array, and preprocessed using the R-package *meffil* (Min et al., 2018), as described in detail elsewhere (Sehovic et al., 2023). EAA estimates, defined as the residuals of chronological age regressed on epigenetic age, were calculated using six different algorithms. Horvath (Horvath, 2013), Hannum (Hannum et al., 2013), PhenoAge (Levine et al., 2018) and GrimAge (Lu et al., 2019) EAA estimates were calculated using their PC-score version (Higgins-Chen et al., 2022). Details about their calculations are described elsewhere (Kankaanpää et al., 2022). In addition, we calculated estimates of EAA with the DunedinPACE (Belsky et al., 2022) and GrimAge2 (Lu et al., 2022) clocks. Although the DunedinPACE algorithm does not strictly correspond to the definition of EAA, it evaluates the pace of biological aging and is therefore comparable to EAA even if its calculation differs. All 642 participants had complete sextuplet of EAA values. EAA estimates showed relatively moderate phenotypic correlations in all twin individuals (mean Pearson correlation: r=0.53; Table S10).

### Proteomics

Proteins from the plasma samples of 786 participants were precipitated and subjected to in-solution digestion according to the standard protocol of the Turku Proteomics Facility (Turku Proteomics Facility, Turku, Finland). Details about protein depletion, precipitation and digestion in this sample have been described elsewhere (Afonin et al., 2024). Samples were first analyzed by independent data acquisition LC-MS/MS using a Q Exactive HF mass spectrometer and further analyzed using Spectronaut software. Data was locally normalized (Callister et al., 2006), and the raw matrix was processed and quality controlled as described elsewhere (Drouard et al., 2023b). Briefly, protein levels were log_2_-transformed and proteins with >10% missing values were excluded. Missing values were imputed by the lowest observed value for each protein carrying missing values. Corrections for batch effects were performed with Combat (Leek et al., 2012) and the final proteomic dataset comprised 439 proteins which were scaled, such that one unit corresponded to one standard deviation (sd). Protein descriptions are available in the supplementary material (Table S1).

### Metabolomics

Metabolites were quantified from plasma samples using high-throughput proton nuclear magnetic resonance spectroscopy (^1^H-NMR) (Nightingale Health Ltd, Helsinki, Finland) (Soininen et al., 2015; Bogl et al., 2016; Rose et al., 2019). Detailed data description (Whipp et al., 2022) are available elsewhere. Data processing included 1) exclusion of pregnant women and individuals taking cholesterol-lowering medications, 2) imputation of missing values using the sample minimum value for metabolites having less than 10% of missing values (NA) and metabolites with >10% missing values were excluded, and 3) examination for the presence of outliers. Metabolites were normalized using inverse normal rank transformation and scaled so that one unit corresponded to a change of one SD, with a mean of zero. The list of metabolites included in the analyses is available in the supplementary material (Table S1).

### External Exposome

The external exposome data comprised a total of 62 exposures derived from Statistics Finland and other sources available for all participants. We used twin’s geocodes in 2005-2006 to merge the exposures, which were described elsewhere (Wang et al., 2023). Briefly, exposures were continuous and included: sizes of and access to green spaces, percentage of built-up areas, population ages and headcounts, crime rates, and voting patterns at municipal elections. Missing values, representing 2.1% of the total data points, were imputed by the median, with the highest missing value rate per variable being 7.5%. A complete description of the exposome’s variables is available in the supplementary material (Table S1).

### Lifestyle

A total of 13 binary variables were selected to describe the lifestyle of the twins, and in particular their education, leisure time activities, substance use, and social behavior. These were derived from questionnaires completed at home and at the study visit when blood samples were taken, as was described elsewhere (Drouard et al., 2023a). Eleven of these variables were initially categorical and represented frequencies: playing video games, watching videos, playing an instrument, reading, going out, dancing, taking part in a club, going to a fast food restaurant, going to a bar, drinking alcohol, and having alcohol-induced blackouts. These variables were dichotomized into binary variables so that the modalities were defined by a frequency of at most once a month vs. at least once a week. Additionally, obtaining a vocational degree (modalities: yes/no) and age of first sexual intercourse (modalities: strictly before age 18/at or after age 18) were included. The list of lifestyle-related variables is available in the supplementary material (Table S1).

We created within-pair, binary lifestyle variables for use in within-pair analyses as well (Drouard et al., 2023a). Briefly, these were coded so that “1” indicated a discordant pair, while “0” indicated a concordant pair. That is, “1” denoted a twin pair in which one twin expressed a feature that the other did not (e.g., one twin often plays video games while the other twin does not), and “0” denoted two co-twins expressing the same trait.

### Covariates

Between-pair analyses included four covariates: sex, body mass index (BMI), and lifetime and current smoking exposure. BMI was calculated from measured height and weight taken during in-person visits at the time of blood sample, and was available for all FinnTwin12 participants except 10; imputation was performed using the median BMI for these 10 participants. BMI ranged from 16.4 to 51.2 kg.m^-2^ (mean: 23.2; SD: 3.9) in the FinnTwin12 sample. Smoking covariates were binary and represented lifetime and current smoking exposure. Lifetime smoking exposure was coded in such a way that twins who had never smoked in their lifetime were coded as ‘1’ and the others as ‘0’. Current smoking exposure was coded so that twins who never smoked, tried smoking but did not smoke, or quit smoking were coded as “1” and compared with twins who currently smoke, coded as “0”. Of the 642 individuals included in the analyses, 102 had never smoked and 249 were current smokers (Table 1).

The same covariates were included in within-pair analyses, namely: sex of the pair, within-pair BMI differences, and lifetime and current smoking discordance. These two latter binary covariates were constructed such that twins within a pair with similar smoking habits (i.e., (“0” ∧ “0”) ∨ (“1” ∧ “1”)) were considered to form a concordant pair, whereas a pair was discordant if two co-twins did not have the same smoking habits (i.e., (“0” ∧ “1”) ∨ (“1” ∧ “0”)). Of the 241 pairs of twins included in the within-pair analyses, 41 and 61 were discordant for lifetime and current smoking exposure, respectively.

## Data processing in external EH-Epi sample

### DNA methylation and EAA calculation

DNA methylation levels were quantified using Infinium Illumina HumanMethylation450K. Data preprocessing is identical to that detailed for the FinnTwin12 sample. A total of 354 participants, including 105 MZ pairs and 72 same-sex DZ pairs, had complete sextuplet EAA values. Correlations between EAA values are in the supplementary material (Table S10).

### Proteomics

Proteomic profiling was initially performed on 415 plasma samples of 120 mL each from participants in the EH-Epi sample. The samples were analyzed using an antibody-based technology (Olink Proteomics AB, Uppsala, Sweden; Olink Explore 384) and processed as described elsewhere (Drouard et al., 2024). Briefly, the data were quality controlled according to Olink’s internal quality control criteria, which led to the rejection of a few outlier samples. Missing values were imputed by the Limit of Detection (LoD) value for proteins with less than 20% missing values; proteins with more missing values were excluded. Final proteomic data were available for 401 EH-Epi twins, of which 354 were included in the current study (Fig.1; Table 1). A total of 2321 proteins were quantified and their values were expressed as Normalized Protein eXpression (NPX) values, where NPX is Olink’s unit for quantifying relative protein levels on a log2 scale. Proteins were then matched by gene name to those in the FinnTwin12 set if they overlapped. Protein descriptions of overlapping proteins used for replication analyses are available in the supplementary material (Table S11).

### Metabolomics

Metabolomic data were generated using high-throughput proton nuclear magnetic resonance (1H-NMR) spectroscopy using the same platform as for the FinnTwin12 sample (Nightingale Health Ltd, Helsinki, Finland). Metabolite values below the limit of quantification were reported as missing, as were metabolite values that deviated from the mean by more than 5 standard deviations (SD). Metabolites with more than 10% missing values were discarded. Missing values (rate of missing values in the dataset: 0.6%) were imputed using the sample minimum of each metabolite for which imputation was to be performed. Outlier presence was assessed by verifying that no participant had at least one of its first three principal components greater than or less than 5 SD from the mean. Metabolite values were normalized using inverse normal rank transformation, with a mean of zero and variance one. In the current study, only metabolites that could be matched to the FinnTwin12 set were used (Table S11).

### Covariates

The use and construction of covariates for between-pair and within-pair analyses was identical to that described for FinnTwin12. BMI was calculated from height and weight measurements taken during in-person visits at the time of blood sample. Description of the EH-Epi sample is shown in Table 1.

### Statistical analyses

Analyses were divided into two main parts. First, between-pair analyses were performed to calculate the associations between EAA and multi-omic factors for all twins. For these analyses, linear mixed-effects models were used. Second, within-pair analyses were performed for all twin pairs, for MZ twin pairs only, and for DZ twin pairs only (Fig.1).

### Between-pair analyses

Linear mixed-effects models were used to quantify the associations between EAA and multi-omic factors. For each EAA variable derived from one of the 6 different algorithms considered, these analyses were performed independently. EAA estimates were modeled as dependent variables. Fixed-effect covariates included sex, BMI, and the two binary variables representing lifetime and current smoking habits (Table 1). To correct for data clustering due to familial relationships among participants, we used family identifiers as a random effect.

The strength of the association between EAA and a multi-omics factor *k* was assessed by testing the nullity of its coefficient β**_k_**. Satterthwaite’s approximation was used to calculate resulting p-values. To counteract the occurrence of false positives, multiple testing corrections were applied. In FinnTwin12 analyses, we corrected the nominal p-values using the number of principal components (PCs) that are needed to cover at least 95% of the initial variance of the omic to which they belong, as calculated by Principal Component Analysis. A nominal p-value was considered significant if it was below α/***Neff***, with α=0.05 and ***Neff*** the number of PCs needed to cover >95% of the original variance. The proteins and metabolites to be replicated in the EH-Epi sample were selected using this last criterion. In the replication analyses conducted in the EH-Epi sample, we adjusted the nominal p-values with Bonferroni correction.

### Within-pair analyses

In within-pair analyses, only complete same-sex twin pairs were included (Fig.1). Analyses were performed on all complete twin pairs, on DZ twin pairs only, and on MZ twin pairs only. The stratification of within-pair analyses by zygosity was motivated by the fact that while the use of all pairs corrects for environmental factors shared between co-twins while maintaining a high level of statistical power, only within-pair analyses using MZ pairs additionally ensure the removal of all genetic confounding in multi-omic associations of EAA. Linear regression models were fitted to model differences in EAA values within a pair, modeled here as the dependent variable, by differences in the multi-omic factors within the pair. Covariates added were sex of the pair, differences in BMI, and lifetime and current smoking binary variables indicating discordance in smoking habits (Table 1). Differences in values were all calculated in such a way that one of the two co-twins was randomly annotated as twin1, from which the value of twin2 was subtracted. This was done in the same order across all datasets. Nullity testing of the coefficient of the multi-omic factor was used to assess the strength of the associations. Correction for multiple testing was performed identically as for between-pair analyses, i.e., correction for effective tests (95% inertia covered by PCs at the omic level) in FinnTwin12 and Bonferroni correction for all associations tested in EH-Epi.

Depending on the degree of genetic confounding affecting the multi-omic associations of EAA, differences in the coefficients obtained in all pairs, DZ pairs only, and MZ pairs only are expected to differ accordingly, as the correction for genetic confounding is either complete in MZ pairs or partial in DZ pairs. Therefore, we compared the coefficients obtained in all pairs (β(within all pairs)), DZ pairs only (β(within DZ pairs)), and MZ pairs only (β(within MZ pairs)). To do so, we notably calculated ratios of the coefficients, such as β(within MZ pairs)/β(within DZ pairs), for which values close to 1 indicate weak genetic effects in the studied association, as the coefficient in MZ twin pairs does not differ substantially to that obtained in DZ twin pairs. Conversely, the lower the β(within MZ pairs)/β(within DZ pairs) ratio, the greater the genetic confounding.

### Data availability

The data analyzed in this study is not publicly available due to the restrictions of informed consent. Requests to access these datasets should be directed to the Institute for Molecular Medicine Finland (FIMM) Data Access Committee (DAC) for authorized researchers who have IRB/ethics approval and an institutionally approved study plan. To ensure the protection of privacy and compliance with national data protection legislation, a data use/transfer agreement is needed, the content and specific clauses of which will depend on the nature of the requested data.

## Acknowledgements

We thank M. Foraster, A. Ambros, and B. Raimbault of ISGlobal for enriching the external exposome data for this study.

## Author contributions

G.D., J.K., and M.O. conceived and designed the study. G.D. and S.S. analyzed the data and performed the statistical analyses. S.S and A.H. preprocessed the methylation data and generated EAA estimates in both FinnTwin12 and EH-Epi samples. Z.W. preprocessed the exposome data of the FinnTwin12 sample. G.D. preprocessed the proteomic, metabolomic and covariate data in both FinnTwin12 and EH-Epi samples, as well as the lifestyle data in the FinnTwin12 sample. M.O. and J.K. contributed to the data collection by providing funding and resources. G.D. wrote the original draft. All authors provided critical feedback on the original draft, participated in its revision, and approved the final version of the manuscript.

## Legends of figures and supplementary tables

**Fig. S1:** Scatter plot of standardized coefficients in both between-pair and within-pair analyses in FinnTwin12.

**Caption:** 37% of the associations in between-pair analyses remained significantly associated with EAA in within-pair analyses in all pairs, MZ-pairs only, or DZ-pairs only (in black). All multi-omic associations that were also significant in the within-pair analyses were of the same direction as those in the between-pair analyses. Golden shaded areas represent zones where the direction of association would not be the same between between-pair and within-pair analyses. Variable descriptions are available in the supplementary material (Table S1).

**Table S1:** Description of multi-omic factors included in between– and within-pair discovery analyses using the FinnTwin12 sample

**Table S2:** Associations between multi-omic factors and epigenetic age acceleration in between-pair analyses using the FinnTwin12 sample

**Table S3:** Repeated between-pair analyses without model adjustment by smoking and body mass index using the FinnTwin12 sample

**Table S4:** Within-pair results in FinnTwin12 using all same-sex twin pairs

**Table S5:** Within-pair results in FinnTwin12 using same-sex dizygotic twin pairs only

**Table S6:** Within-pair results in FinnTwin12 using monozygotic twin pairs only

**Table S7:** Within-pair coefficients in MZ and DZ pairs separately for associations found in within-pair models using all pairs

**Table S8:** Associations between multi-omic factors and epigenetic age acceleration in between-pair analyses using the EH-Epi replication sample

**Table S9:** Replication of within-pair results from FinnTwin12 in external EH-Epi sample in all same-sex twin pairs, same-sex DZ pairs only and MZ pairs only

**Table S10:** Phenotypic correlations between EAA estimates in FinnTwin12 twin individuals – why not EH-Epi too

**Table S11:** Description of multi-omic factors included in between– and within-pair replication analyses using the EH-Epi sample

