## Supplementary figures and images for "Multi-omic associations of epigenetic age acceleration are heterogeneously shaped by genetic and environmental influences"

### Fig.S1

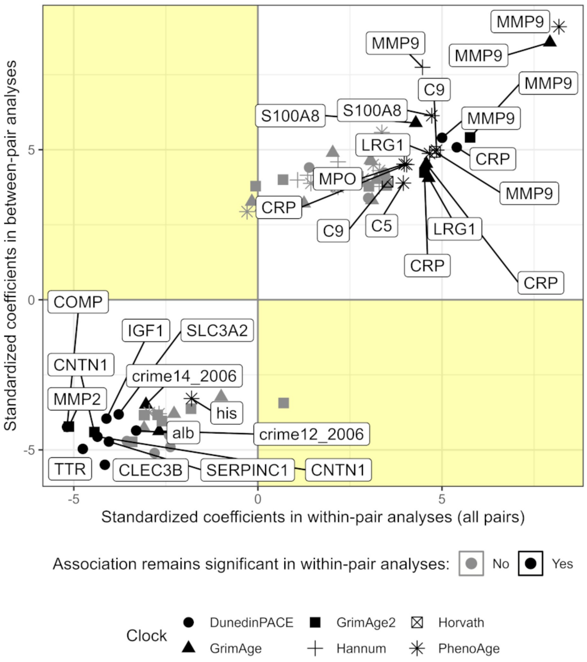
